# Word embeddings trained on published case reports are lightweight, effective for clinical tasks, and free of protected health information

**DOI:** 10.1101/19013268

**Authors:** Zachary N. Flamholz, Lyle H. Ungar, Gary E. Weissman

**Affiliations:** Medical Scientist Training Program (MSTP), Albert Einstein College of Medicine, Bronx, New York; Department of Computer and Information Science, University of Pennsylvania, Philadelphia, Pennsylvania; Department of Medicine, Perelman School of Medicine, University of Pennsylvania, Philadelphia, Pennsylvania

## Abstract

**Rationale:** Word embeddings are used to create vector representations of text data but not all embeddings appropriately capture clinical information, are free of protected health information, and are computationally accessible to most researchers.

**Methods:** We trained word embeddings on published case reports because their language mimics that of clinical notes, the manuscripts are already de-identified by virtue of being published, and the corpus is much smaller than those trained on large, publicly available datasets. We tested the performance of these embeddings across five clinically relevant tasks and compared the results to embeddings trained on a large Wikipedia corpus, all publicly available manuscripts, notes from the MIMIC-III database using fastText, GloVe, and word2vec, and using different dimensions. Tasks included clinical applications of lexicographic coverage, semantic similarity, clustering purity, linguistic regularity, and mortality prediction.

**Results:** The embeddings trained using the published case reports performed as well as if not better on most tasks than those using other corpora. The embeddings trained using all published manuscripts had the most consistent performance across all tasks and required a corpus with 100 times as many tokens as the corpus comprised of only case reports. Embeddings trained on the MIMIC-III dataset had small but marginally better scores on the clustering tasks which was also based on clinical notes from the MIMIC-III dataset. Embeddings trained on the Wikipedia corpus, although containing almost twice as many tokens as all available published manuscripts, performed poorly compared to those trained on medical and clinical corpora.

**Conclusion:** Word embeddings trained on freely available published case reports performed well for most clinical task, are free of protected health information, and are small compared to commonly used embeddings trained on larger clinical and non-clinical corpora. The optimal corpus, dimension size, and which embedding model to use for a given task involves tradeoffs in privacy, reproducibility, performance, and computational resources.

## Introduction

Word embeddings are a low-dimensional, vector representation of semantic meaning that permit efficient analysis of text data. While such embeddings are useful across many domains, their application in analyzing the text of clinical encounter notes presents several challenges. First, commonly available pre-trained embeddings are derived from a broad range of non-medical text sources and thus may not capture the appropriate word sense, linguistic relations, or vocabulary needed in a clinical context [1]. Second, while it may be appealing to train new embeddings on locally available clincally notes within a health system [2], clincal corpora from a single center may not be sufficiently large compared to publicly available corpora [3], and such embeddings cannot be shared because they would contain protected health information, thus limiting the reproducibility of experiments that would rely on such representations.

Prior work on domain-specific word embeddings has focused on knowledge discovery in large corpora of biomedical text [4], using medically-relevant articles from Wikipedia [5] and MEDLINE abstracts [6], and using the MIMIC-III dataset which is de-identified, but requires a data use agreement for researchers and is limited to a single center [7]. Other work has focused on concept embeddings [8] that capture clinical information from both structured and unstructured data sources and relies on pre-processing with the Unified Medical Language System (UMLS) thesaurus. None of these approaches has yet to provide a robust evaluation of a set of embeddings specific to the text of clinical encounter notes that is lightweight, free of any protected health information, and readily shareable with other researchers as a stand-alone tool.

To overcome these barriers, we developed a training corpus from the clinical case reports in the PubMed Open Access subset. We hypothesized that 1) embeddings trained on the full text of clinical case reports (which are already fully de-identified as a result of the editorial and publication process) would share similar lexical and syntactic properties with clinical notes and outperform embeddings from a non-medical domain; and 2) that subword n-grams [9] would outperform word-level n-grams [10] due to their ability to produce word vectors for out-of-vocabulary and misspelled terms, which occur frequently in electronic health record data.

## Methods

We trained a set of word embeddings using every combination of three different model types, four different corpora, and four different dimension sizes (Table 1). We then tested each set of embeddings with five clinical tasks. All models trained on the Open Access Subset are available for free download: https://github.com/weissman-lab/clinical_embeddings

**Table 1:** Summary of model types, corpora, and dimensions used to train word embeddings.

| Model Type | Corpus | Dimensions |
| --- | --- | --- |
| word2vec | PMC Open Access Subset – All manuscripts (OA-All) | 100 |
| fastText | PMC Open Access Subset – Case reports only (OA-CR) | 300 |
| GloVe | MIMIC-III (MIMIC) | 600 |
|  | Wikipedia – English (Wiki) | 1200 |

### Word Embedding Models

We trained embeddings using word2vec [10] and fastText [9] with a skip-gram architecture as implemented in the Python gensim package [11]. We trained GloVe embeddings using GloVe software (version 1.2) [12]. Embeddings were built with 100, 300, 600, and 1200 dimensions.

All models trained with gensim used the following hyperparameters: skip-gram (sg=1), window=7, min_count=5, sorted_vocab=1, seed=2018. The fastText models used the additional hyperparameters: word_ngrams=1, min_n=3, max_n=8. Hyperparameters for GloVe models included: VOCAB_MIN_COUNT=5, WINDOW_SIZE=7, MAX_ITER=15.

### Text pre-processing

For the OA-CR and MIMIC corpora, the entire corpus was first tokenized by sentence and then by word using the Spacy tokenizer [13]. For the longer OA-ALL and Wiki corpora, the gensim `LineSentence` method was used to tokenize the corpus for input into the models. Corpora were saved as single text files for input into the GloVe model. For all corpora, all years were normalized to a single token “year”; then all real numbers were normalized to “real_number”“; and all integers were normalized to”integer”. We did not remove stopwords or perform stemming.

### Corpus selection and corpus-specific pre-processing

#### PMC Open Access Subset

The PubMed Open Access Case Reports (OA-CR) corpus was built using case report manuscripts downloaded from PubMed Central with the query `“Case Reports[ptyp] AND “2007/01/01”[PDat] : “2017/12/31”[PDat] AND English [lang] AND “humans”[MeSH Terms]”` that were available under the OpenAccess Subset as identified using the “oa_file_list.csv” (ftp://ftp.ncbi.nlm.nih.gov/pub/pmc/oa_file_list.csv). Of the 515,592 reports returned by the original query, 27,575 were openly available. Text was extracted from the downloaded XML files from the abstract and body sections. All non-English text was removed using the detect method of the `langdetect` python package [14]. All text was converted to lowercase and stripped of non-body text, XML tags, breaks and tabs, figure tables and captions, figure references, citations, and URLs. Reports with less than 100 tokens of processed text were removed.

The PubMed OpenAccess all reports (OA-All) corpus was built using all manuscripts downloaded from PubMed Central with the query `“2007/01/01”[PDat] : “2017/12/31”[PDat] AND English[lang] AND “humans”[MeSH Terms]` that were available under the OpenAccess Subset as described above. Of the 5,834,856 reports returned by the original query, 630,885 were openly available. XML files were processed exactly as described for the OA-CR corpus.

#### MIMIC

The MIMIC clinical notes (MIMIC) corpus was built using the NOTEEVENTS.csv file in the MIMIC-III v1.4 dataset [15,16]. 278,269 notes were extracted and processed. All text was converted to lowercase and stripped of end fields, generic fields, de-identified notation, underscores used as separation lines, and breaks and tabs. Documents with less than 50 tokens of processed text were removed.

#### Wikipedia

The Wikipedia (Wiki) corpus was built using a Wikipedia dump (downloaded 2018-11-02, 15.6GB). The download contained 18,906,413 articles, which were processed using the WikiCorpus method from the gensim python package [11]. Articles with less than 50 words were removed.

#### Multi-word expressions

Multiword expressions were identified using both an intrinsic and extrinsic method. Intrinsically, pointwise mutual information (PMI) was calculated for every bigram and trigram in the OA-CR corpus using the nltk python package [17]. Bigrams and trigrams were included if they appeared in at least 10 unique manuscripts. Bigrams and trigrams scoring in the 50th to 95th percentile of PMI were kept based on our manual review of clinical relevance. Extrinsically, the National Library of Medicine specialist lexicon [18] was used to identify MWEs by the presence of lexicon terms in the OA-CR corpus. In total, 398,217 n-grams were classified as MWEs. The `KeywordProcessor` method in the `flashtext` python package [19] was used to join all MWEs with an underscore between words in all corpora for training embeddings and for all tasks.

### Tasks

#### Lexicographic Coverage

Models were evaluated on their ability to provide a vector representations for all the words in a given text. A useful set of word embeddings should be able to represent a wide variety of medical terms, including new terms that might not have been seen in an original training corpus, and misspellings in the electronic health record. We tested coverage using all 362,430 unique words in 53,425 MIMIC ICU discharge summary notes [15,16]. Coverage is reported as the proportion of all tokens for which a model could provide a word vector.

#### Semantic Similarity

Semantic similarity was measured by computing the correlation between the cosine distance between word pairs and the manually curated similarity scores of those pairs in the UMNSRS similarity dataset [20]. This tasks measures the degree to which vectorized word meanings correspond to a human-annotated, clinical sense of the word. Correlations are reported using Spearman’s *ρ*. Only word pairs for which both terms had a vector representation were considered in the comparison.

#### Clustering Purity

Clustering purity was measured by creating a document-level vector representation of the discharge summaries from six different ICUs in the MIMIC dataset. Each document was represented by calculating the centroid across all individual word vectors. Words for which no vector was available were ignored. Duplicate reports and addenda were removed, leaving 49,698 ICU discharge notes. A K-means procedure (*k* = 6) was used to cluster the document-level vectors. Clustering purity was calculated by summing the number of correctly assigned notes, where ICU type is assigned to a class based on the ICU type that received the maximum number of notes for that class, and dividing by the total number of notes. Additionally, we report the 95% bootstrapped confidence interval of 1,000 replicates for the purity measure.

#### Linguistic Regularity

A known feature of continuous space language models is the preservation of an offset vector that captures some semantic regularity [21]. A useful set of clinically-relevant word embeddings should capture offset vectors related to medical treatment and using appropriate medical terminology. We curated a list of 100 pairs of medical terms with a relationship `is_a_treatment_for` across inpatient, outpatient, medical, and surgical contexts likely to be discussed in a clinical encounter note. For example, metformin is a treatment for diabetes just as lisinopril is a treatment for hypertension. Therefore, an embedding that captures clinically relevant semantic information should reproduce the analogy: *v*_*metformin*_ − *v*_*diabetes*_ = *v*_*lisinopril*_ − *v*_*hypertension*_.

We used two approaches to measure how well this treatment relationship was preserved in the embedding space for each model. First, we computed the vector difference for each pair of 100 terms that was preselected to have a `is_a_treatment_for` relationship, then computed the cosine similarity between that and the centroid of all vectors. The standard deviation of this set of similarities is reported as a measure of the regularity of this clinical relationship. Embeddings with low measures of variance have a more regular representation of the treatment relationship in these pairs.

Second, we used a previously reported analogy completion task [22]. For every combination of pairs, representing an analogy *a* : *b* :: *c* : *d*, we calculated the cosine similarity between *d* and the single closest vocabulary word in the embedding space to *a* − *b* + *c*.

#### Mortality Prediction

We also tested each set of embeddings by using their respective vector representations of the text of clinical notes as inputs to a mortality prediction model. The first physician encounter note charted within 24 hours of hospital admission in the MIMIC dataset was used to predict in-hospital mortality using a convolutional neural network. The network architecture was comprised of an input layer, a convolutional layer, a max pooling layer, a dense layer with 128 nodes, and an output layer with a single node using a sigmoid activation. Of the 4,170 notes, 522 (12.5%) were associated with a patient death during the hospitalization. The collection of notes was randomly split into 80% and 20% groups for training and testing, respectively. Sampling was stratified to maintain balance in the outcome between the two sets. We reported performance of each model using the mean Brier score over 1,000 runs with a 95% confidence interval on the testing sample.

## Results

The Wikipedia corpus was the largest and contained more than 100 times the number of tokens as the Open Access Case Reports corpus (Table 2). Although the MIMIC dataset contained almost ten times as many tokens as the Open Access Case Reports, the number of unique words identified for the embeddings was about half across all model types.

**Table 2:** Summary of corpora.

| Corpus | Documents | Tokens | Words - Word2Vec | Words - fastText | Words - GloVe |
| --- | --- | --- | --- | --- | --- |
| Open Access Case Reports | 27,449 | 49,590,835 | 333,360 | 333,360 | 435,835 |
| MIMIC-III | 220,453 | 148,089,760 | 160,411 | 160,411 | 267,629 |
| Wikipedia | 4,555,827 | 2,542,552,916 | 3,338,426 | 3,338,426 | 3,338,427 |
| Open Access - All | 628,404 | 1,848,856,520 | 3,748,342 | 3,748,342 | 3,755,370 |

### Lexicographic Coverage

The fastText models had perfect coverage (Table 3), as embeddings for out-of-vocabulary words are built from character-level n-grams. Although the total vocabulary for the GloVe models was larger, all word2vec models had as much if not greater coverage.

**Table 3:**
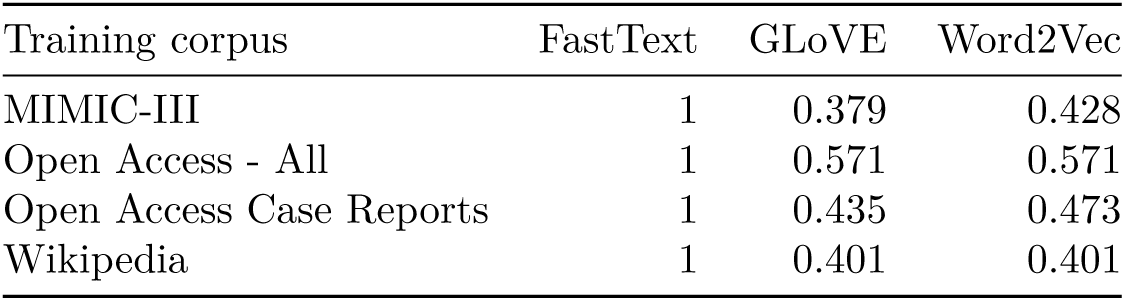
Lexical coverage for each model measured as the proportion of 362,430 unique tokens in 53,425 MIMIC ICU discharge summary notes for which each model could produce a word vector.

### Semantic Similarity

Models trained on the smaller clinical corpora consistently outperformed those trained on the larger Wikipedia corpus (Figure 1). Word2vec models outperformed FastText models with all corpora except in the higher dimension Wiki models. GloVe models consistently performed worst, except in the Wiki models where the higher dimension models scored the best for the corpus.

**Figure 1:**
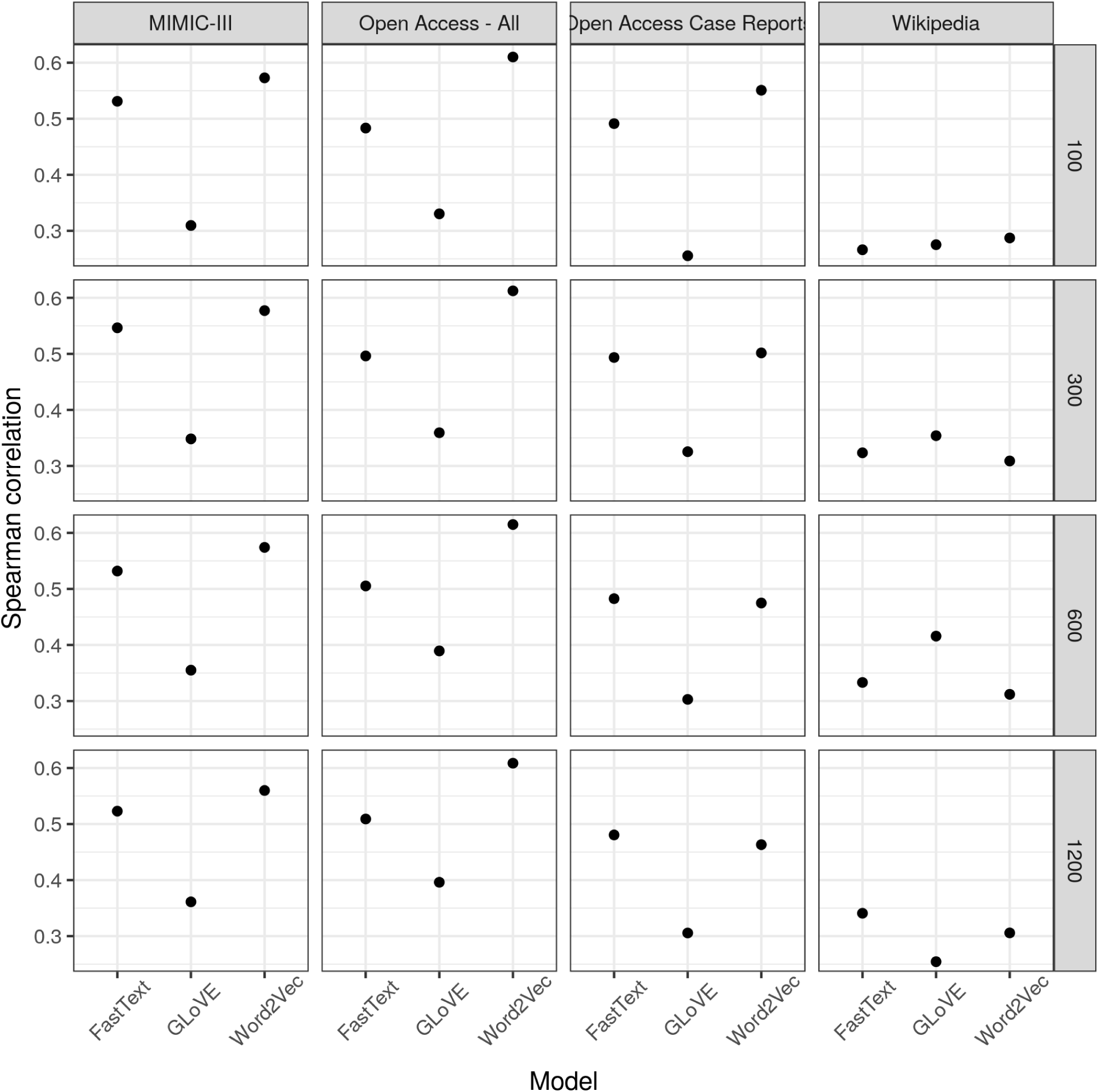
Spearman correlation between the cosine similarity of the words in each pair and the manually annotated similarity.

### Clustering Purity

The embeddings trained with fastText produced the highest clustering purity across nearly all categories (Figure 2). With the exception of the GloVe models, which performed poorly for this task, those models trained on clinical corpora outperformed those trained with the Wikipedia corpus. Orthographically similar words in fastText embeddings appeared to be more tightly clustered than those trained with word2vec or GloVe (Figure 3).

**Figure 2:**
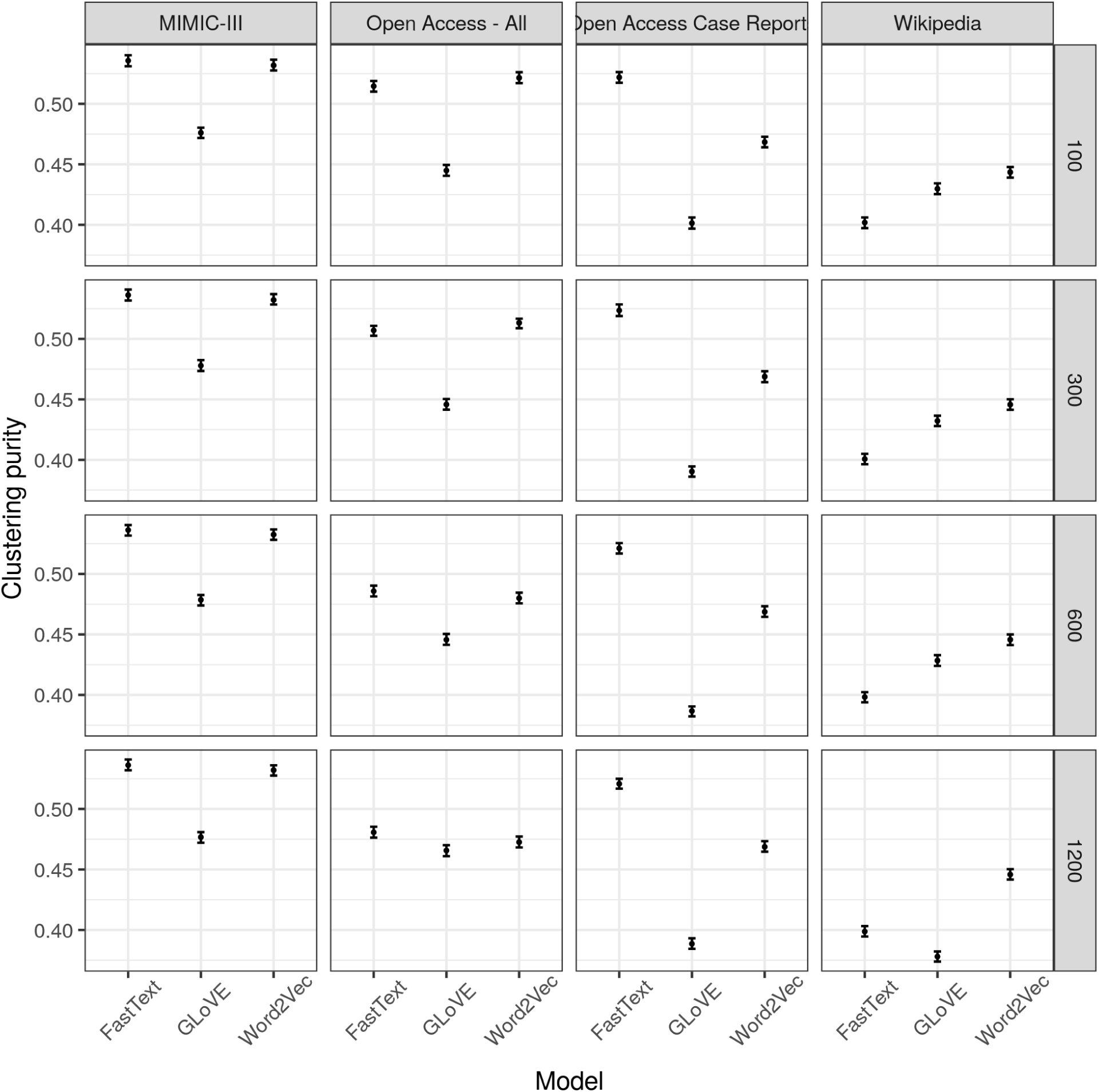
Clustering purity based on a k-means procedure using document-level vectors for discharge summaries from six intensive care units in the MIMIC-III dataset.

**Figure 3:**
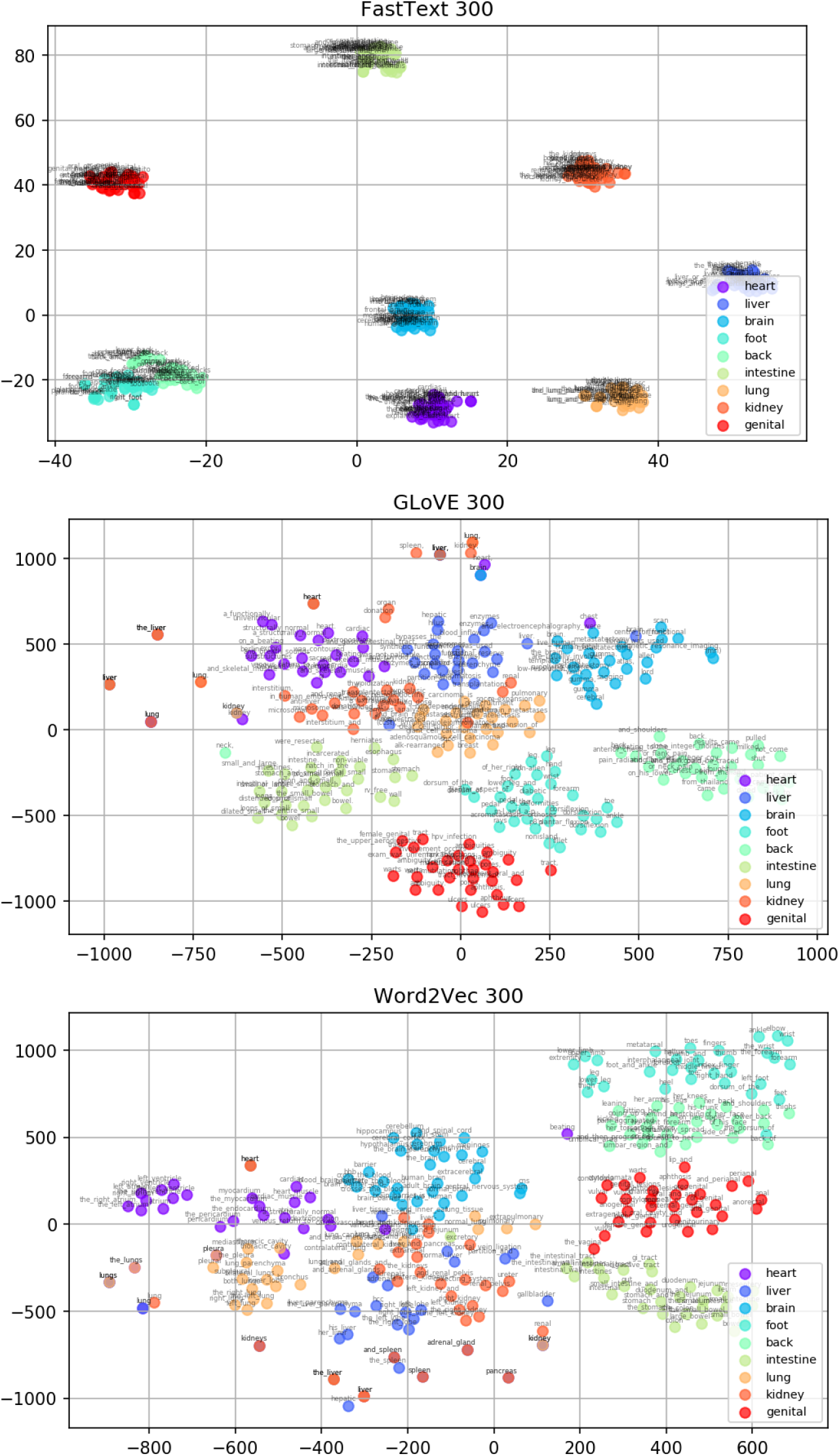
tSNE plot of 300-dimensional embeddings trained with clinical case reports from the PubMed Open Access Subset using fastText (top), GloVe (middle), and word2vec (bottom).

### Linguistic Regularity

Models trained with the Open Access Case report corpus had the best performance, followed by those trained with all Open Access manuscripts (Figures 4, 5.

**Figure 4:**
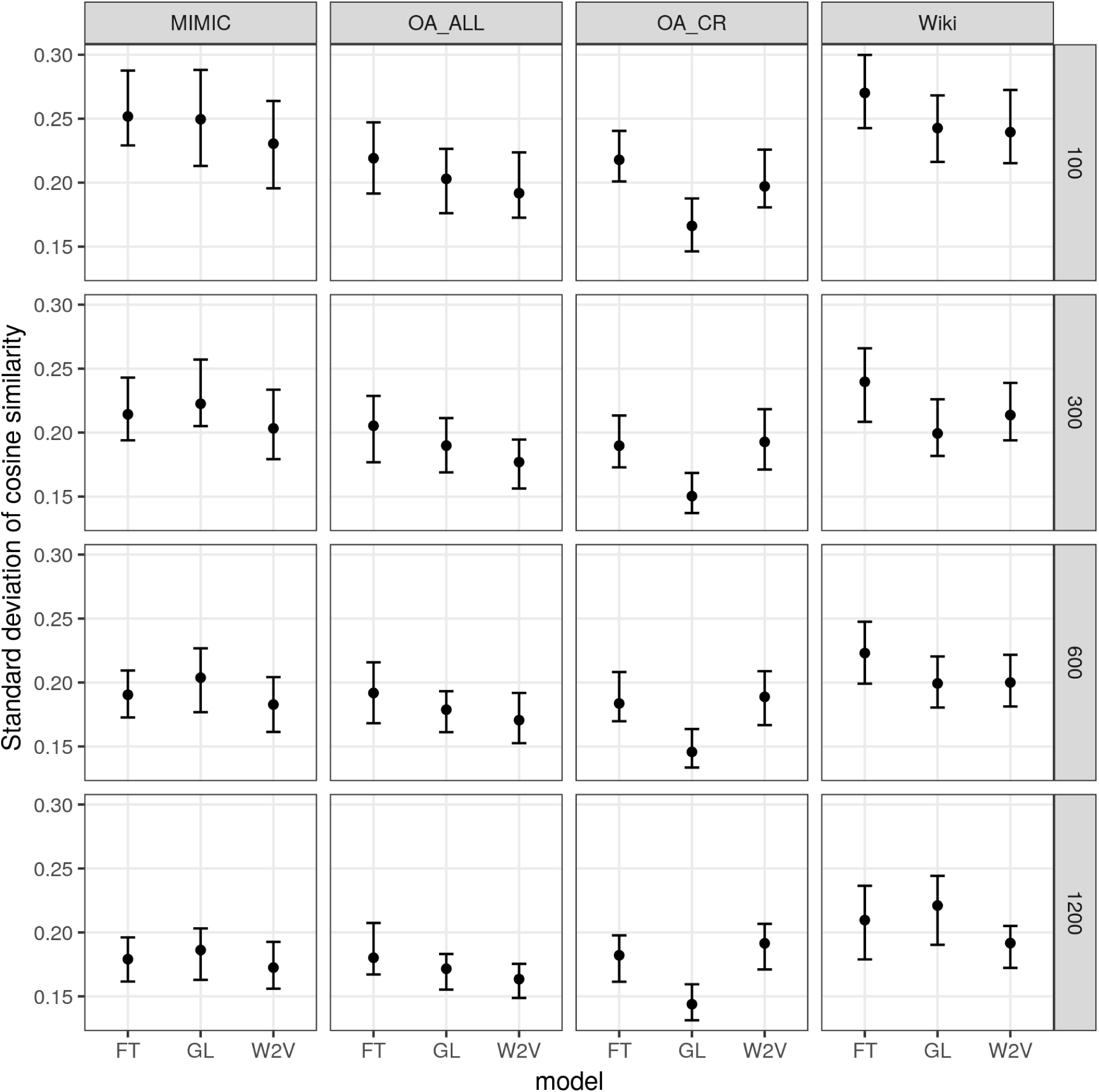
The variance in cosine similarity across vector differences of 100 word pairs related by `is_a_treatment_for`. Embeddings with lower standard deviation capture a more regular treatment relationship using the same vector difference.

**Figure 5:**
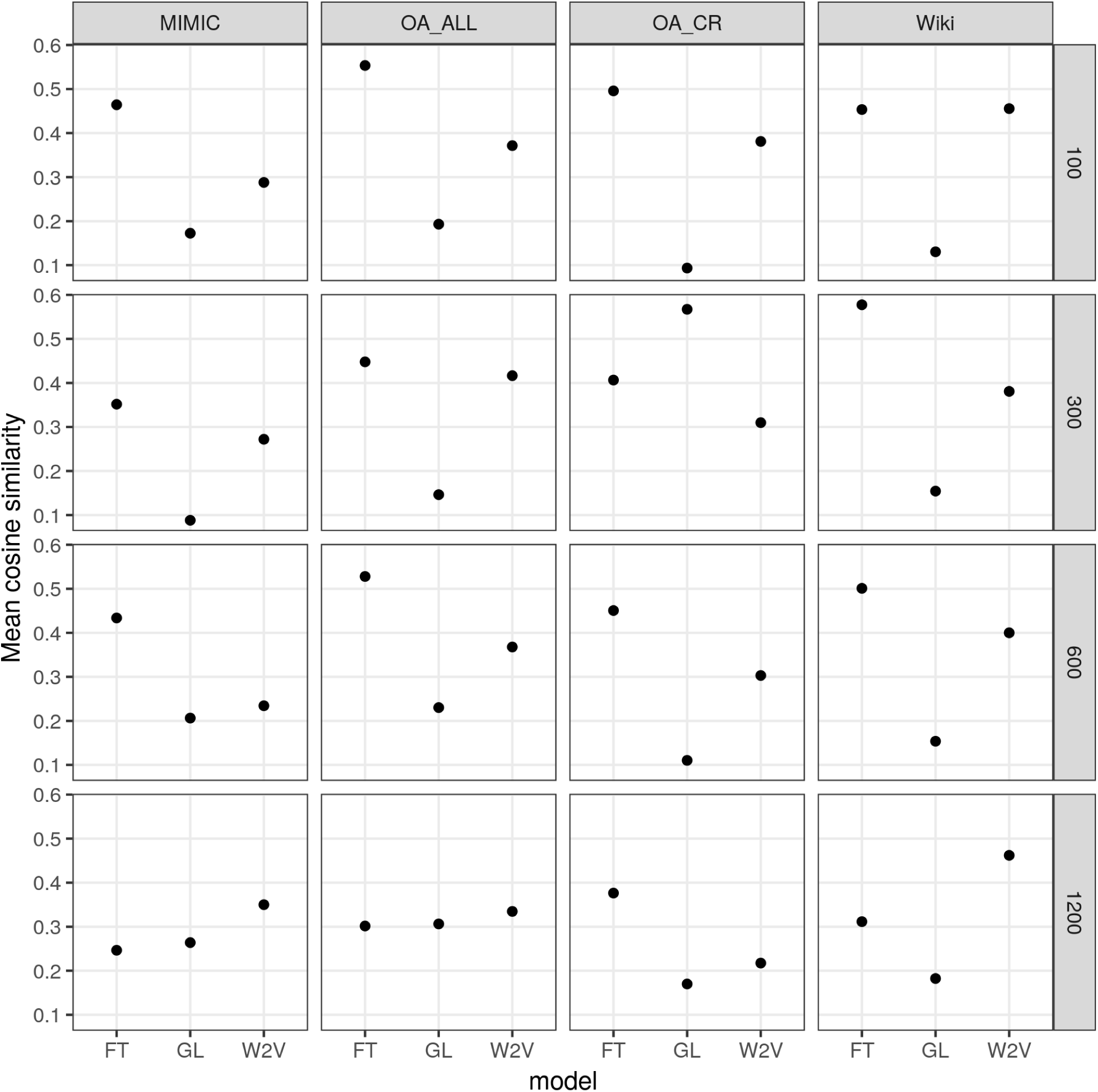
Mean cosine similarity between *a* − *b* + *c* and *d* across every combination of 100 pairs of treatment:disease (4,950 analogies).

### Mortality Prediction

Models using vector inputs derived from clinical corpora performed better across higher dimensions (Figure 6). The word2vec model with 300 dimensions trained on all OA manuscripts had the best performance (Brier Score 0.13). Models trained on Wikipedia performed best only at 100 dimensions. At a dimension size of 100, Wiki models outperformed the other corpora, but it performed worse for higher dimensions. Both OA-CR and OA-All Word2Vec models outperformed MIMIC Word2Vec models, but not fastText models. GloVe models performed worst across all corpora.

**Figure 6:**
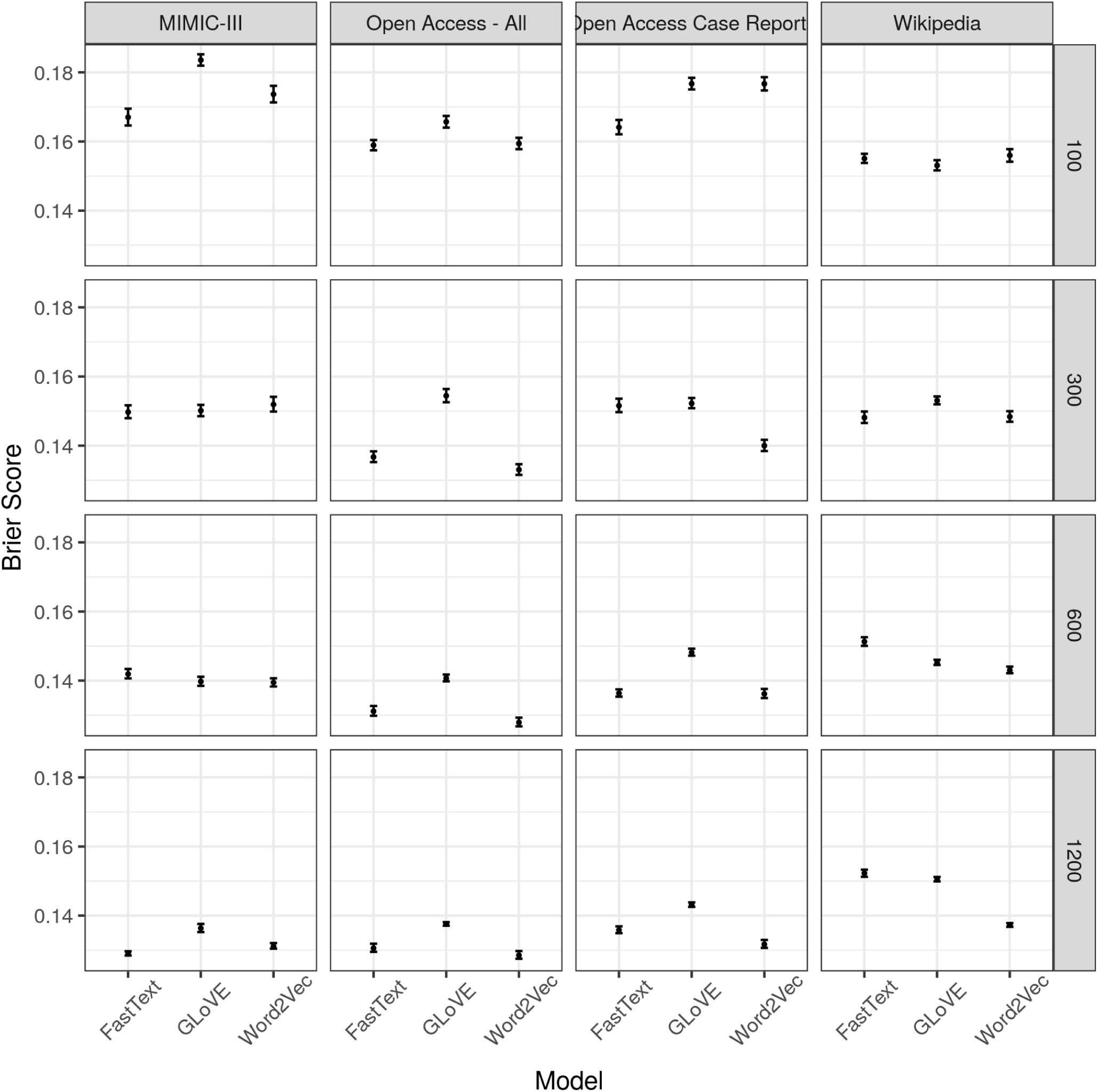
Performance of a mortality prediction model using only the first physician note in the first 24 hours of hospitalization. Performance is reported on the testing set using the Brier Score.

## Discussion

Word embeddings trained on the OA-CR corpus led to performance on par with, and often better than, embeddings trained on larger text corpora in a number of semantic and other NLP tasks. Optimal performance in a specific task, however, varied by corpus and model. The ability to learn word meanings and relationships relevant in the clinical domain is a major advantage in the biomedical informatics community, and while it has been shown that embeddings trained on clinical-domain-specific text perform better than general-language corpora [2], we show that the lightweight embeddings trained on publically available case reports can be effective in clinically relevant NLP tasks.

Our general language corpus, Wiki, was the larget corpis in terms of both documents and words. However, it performed worst in the lexicographic coverage task, scoring lower than even the OA-CR models, which were built on a corpus that was two orders of magnitude smaller. Though the Wiki corpus was the largest, the OA-All embedding models contained more words. This may be explained by the requirement that a word must appear at least five times in a corpus to be included in the model. An obscure medical term that appears in a manuscript is more likely to appear at least five times than an obscure word in a Wikipedia article that may have fewer mentions. The OA-All models performed the best on the lexicographic coverage task. Finally, we note that the GLoVE OA-CR and MIMIC models have more words in them but perfom worse on the lexicographic coverage task. This may be explained by the use of the same tokenizer in preparing the Word2Vec and FastText models and preparing the coverage test dataset, which was not used in preparing the GLoVE models.

We used the MIMIC-III clinical notes as a proxy for a local, health-system based clinical text corpus. The MIMIC corpus size is smaller than other clinical corpora used [1]. Additionally, it is important to note that the lexicographic coverage, ICU clustering, and mortality prediction tasks all utilized MIMIC-III notes for testing. The parallel performance of the models built using OpenAccess text highlight the versatility of those models in capturing clinical encounter text.

Our findings also highlight several opportunities for future work. In the UMNSRS semantic relatedness task, our highest performing model performed as well as the highest performing model reported by [1], which was also built from manuscripts in the PMC database but was not limited to those in the OpenAccess subset. Additionally, the performance of the models in the MIMIC-III based mortality prediction task is better than the performance reported in mortality prediction of severe sepsis patients [23], but worse than the performance reported in acute kidney injury patients [24]. However, our models made use solely of the text of the admission note, and could be improved by the inclusion of other data points related to the admission.

Overall, the GLoVE models performed worse than the Word2Vec and FastText models, and the Wiki corpus models performed worst among the corpora. Embedding dimension size did not affect the results of the semantic relatedness and clustering purity tasks. It did have an effect on mortality prediction. The effect of dimension size on the linguistic regularity task was ambiguous, however, the observed effect on the standard deviation measure could be the result of the increased embedding space.

In visualizing the embeddings there is a stark contrast between the tight clusters of the FastText model and the more dispersed clusters of the Word2Vec and GLoVE models. Looking at the words that cluster around a term it was evident that the FastText clusters were comprised of words in which the term was a subword of the word, for example “heart” produced “hearth” and “whole_heart”, while the other models did not produce these words. As FastText learns embeddings for n-grams of words, the subword will learn every context of all the words of which it is a subword. We hypothesize that this sharing of contexts place the words in close proximity in the embedding space. This also represents a limitation of our multiword expression procedure, as a single word will always be an n-gram of the joined multiword expression.

In this study we show that embeddings trained on manuscripts from the PMC OpenAccess database capture clinically relevant word meanings, and that a substantially smaller subset made up of only case reports can perform very well for clinical NLP tasks. We evaluated the models for their ability to capture clinical meaning and be used for downstream predictive tasks. By outlining how we built our models and measuring their performance in a variety of ways, we hope to make word embeddings more accessible to the broader clinical research informatics community. Furthermore, by evaluating FastText, GLoVE, and Word2Vec, as well as for multiple dimension sizes, we hope that our results serve as a benchmark for embeddings built from other corpora.

## Data Availability

Data are all publicly available. Code and final word embedding are available for free download from the author's GitHub page.

https://github.com/gweissman/clinical_embeddings

